# Linking microstructural integrity and motor cortex excitability in multiple sclerosis

**DOI:** 10.1101/2020.10.15.20213090

**Authors:** Angela Radetz, Kalina Mladenova, Dumitru Ciolac, Gabriel Gonzalez-Escamilla, Vinzenz Fleischer, Erik Ellwardt, Julia Krämer, Stefan Bittner, Sven G. Meuth, Muthuraman Muthuraman, Sergiu Groppa

## Abstract

Motor skills are frequently impaired in multiple sclerosis (MS) patients following grey (GM) and white matter (WM) damage with cortical excitability abnormalities. We performed advanced diffusion imaging for neurite orientation dispersion and density modeling and diffusion tensor imaging within the motor system of 50 MS patients and 49 age-matched healthy controls. To assess excitability, we determined resting motor thresholds using non-invasive transcranial magnetic stimulation. A hierarchical regression model revealed that lower neurite density index (NDI), suggestive for axonal loss in the GM, predicted higher motor thresholds, i.e. reduced excitability in MS patients. Furthermore, lower NDI was indicative of decreased cognitive-motor performance. Interconnected motor WM tracts of patients were characterized by overlapping clusters of lowered fractional anisotropy and NDI, with NDI exclusively capturing a higher amount of abnormally appearing voxels. Our work outlines the potential of microstructure imaging using advanced biophysical models to forecast neurodegeneration and excitability alterations in neuroinflammation.

## Introduction

Impaired motor functions caused by both grey (GM) and white matter (WM) pathology are characteristic of multiple sclerosis (MS), a neurodegenerative inflammatory disorder of the central nervous system (Eshaghi et al., 2018; Filippi et al., 2012; Kurtzke, 1983). Both in animal models of MS and in patients, cortical neurons show abnormal levels of excitability, indicating a pathophysiological relevance in neuroinflammation (Caramia et al., 2004; Ellwardt et al., 2018). Excitability of neural tissue in motor brain regions can be measured in-vivo by determining the resting motor threshold with transcranial magnetic stimulation (TMS). Hereby, increased resting motor thresholds were frequently observed in MS patients (Stampanoni Bassi et al., 2020; Zipser et al., 2018). Although the driving mechanisms of altered cortical excitability are still unclear, cortical neuronal or axonal loss or altered (re-) myelination processes could play an important role (Calabrese et al., 2007; Zipser et al., 2018). Indeed, GM integrity alterations are a pathophysiological signature of MS; atrophy quantified with conventional imaging sequences is commonly used to track disease courses (Eshaghi et al., 2018; Filippi et al., 2012; Radetz et al., 2019). Whereas this measure does not allow drawing conclusions about the depicted microstructure, non-conventional imaging approaches allow an in-vivo quantification of microstructural alterations caused by neurodegenerative processes (Alexander et al., 2019). Monitoring and assessing microstructural alterations might improve the understanding of altered excitability levels. With diffusion-weighted imaging (DWI) sequences, WM microstructural integrity has been robustly quantified based on parameters derived from fitted diffusion tensors (diffusion tensor imaging [DTI]), such as fractional anisotropy (FA) (Deppe, Krämer, et al., 2016; Deppe, Tabelow, et al., 2016; Droby et al., 2015; Roosendaal et al., 2009). Whereas FA is generally sensitive to microstructural neuroinflammation-or neurodegeneration-driven tissue damage, it is relatively non-specific for the underlying pathophysiological processes (Krämer et al., 2019). FA does not distinguish restricted and hindered diffusion and is thereby biased in areas with high neurite orientation dispersion such as the GM or regions of crossing fibers (Tournier et al., 2011; Zhang et al., 2012). Advanced biophysical diffusion models that are continuously refined allow a more realistic in-vivo depiction of GM and WM microstructure including estimations of neurite density, axon diameter, neurite dispersion and cell size (Alexander et al., 2019; Lakhani et al., 2020). The neurite orientation dispersion and density imaging (NODDI) model, which is based on advanced acquisition techniques, mitigates the methodological constraints as described for FA (Zhang et al., 2012). In each voxel a neurite density (NDI) and orientation dispersion index (ODI) is computed from restricted diffusion, whereas an isotropic volume fraction (IVF) reflects cerebrospinal fluid and edema (Zhang et al., 2012). A mouse diffusion imaging study showed good agreement of the model with parallel histological quantification of microstructure. High ODI values were reported in areas of complex architecture or crossing fibers, and high NDI values in regions with low neuron content and dense fiber bundles (Wang et al., 2019). A cross-modality study of the human cortical GM suggested a strong link between NDI and the density of myelinated, and to a lesser extent of unmyelinated axons. ODI was proposed to reflect cyto- and myeloarchitecture (Fukutomi et al., 2018). Importantly, both ODI and NDI contribute to FA, thereby allowing a more specific differentiation of microstructural characteristics (Zhang et al., 2012). Here, we aimed at carrying forward the potential of the NODDI model to evaluate the microstructural integrity in the GM of primary motor cortex and address the question, whether lower excitability is related to pathological changes characterized by NDI, ODI or IVF. We further hypothesized that NODDI measures used for quantification of motor microstructural integrity are predictive for motor and cognitive function. Such assessments are of pivotal importance as the applied clinically feasible diffusion sequence could particularly forecast neurodegenerative processes that lead to progression independent of relapse activity.

As neuroinflammation with de- and remyelination, as well as neurodegeneration occurs in GM and WM, we emphasize the importance of gaining a robust depiction of GM and WM pathology specifically of the motor system for the prediction of the functional trajectories. At the whole-brain level, reduced NDI in the normal-appearing WM of MS patients compared to that of healthy controls (HC) was previously reported, indicating myelin and axonal loss (Granberg et al., 2017; Spano et al., 2018). ODI generally yielded more variable results during comparisons between MS and HC (De Santis et al., 2019; Lakhani et al., 2020; Spano et al., 2018). Alterations in ODI result from morphological changes including a combination of axonal loss, dendritic pruning and tissue reorganization. Therefore, conclusions about the pathological processes strongly depend on the original tissue architecture (Spano et al., 2018). We here compared microstructural integrity in motor WM tracts of MS patients and HC using tract-based spatial statistics (TBSS) (Smith et al., 2006). This method allows for voxel-wise group comparisons of FA and NODDI parameters in WM tracts by minimizing partial volume effects. Importantly, we investigated if NODDI values in contrast to the classical FA are more sensitive in detecting differences in microstructural integrity in MS patients versus HC. As FA increases might reflect either NDI increases, ODI decreases or a combination of both (Zhang et al., 2012), we tracked contributions of these signals to FA and evaluated if the NODDI model captures additional pathological alterations due to MS pathology. This approach allows more specific inferences about the type of microstructural pathology underlying the FA changes.

We first assessed the predictive value of motor cortex microstructure for motor excitability and cognitive-motor performance in our participants. Next, we evaluated pathological microstructure alterations in motor WM tracts of MS patients and HC using NODDI in comparison to the classical FA. We hereby examined voxel-wise intersecting group differences to disentangle potential contributions of NODDI metrics to FA.

## Methods

### Participants

Fifty patients with relapsing-remitting MS (RRMS) (31 female, mean age = 35.3 years, SD = 11.1), diagnosed according to the revised McDonald criteria (Thompson et al., 2018), and 49 HC (25 female, mean age = 31.5 years, SD = 9.1) participated in our study. Clinical data of patients, Nine Hole Peg Test (9HPT), Trail Making Test part A (TMT-A) and part B (TMT-B) raw scores (Reitan, 1971) and an overview of ongoing therapies at the time of MRI acquisition are depicted in Table 1. Participants were excluded from the study in case of pregnancy or relapses or systemic therapy with steroids within the month before MRI. They were also excluded in case of contraindication to TMS, i.e. previous or current susceptibility to epileptic seizures or metal implants in proximity to the cranium. Expanded Disability Status Scale (EDSS) scores (Kurtzke, 1983) of MS patients were determined by trained and certified neurologists. The study was approved by the local Ethics Committee of the State Medical Association of Rhineland-Palatinate. All participants gave written informed consent before participation and were informed about the study content in oral and written form.

**Table 1.** Overview of patients’ clinical characteristics

|  |  |
| --- | --- |
| Disease duration in months, mean ± SD | 50 ± 59 |
| EDSS, median (interquartile range) | 1 (1.25) |
| Total lesion volume in ml, mean ± SD | 3.88 ± 5.98 |
| 9HPT raw score, mean ± SD | 67.47 ± 13.48 |
| TMT-A raw score, mean ± SD | 25.40 ± 11.82 |
| TMT-B raw score, mean ± SD | 52.50 ± 28.04 |
| Disease modifying therapies |  |
| Dimethyl fumarate | 14 |
| Beta interferons | 10 |
| Glatiramer acetate | 5 |
| Natalizumab | 5 |
| Fingolimod | 3 |
| Alemtuzumab | 2 |
| Daclizumab | 2 |
| Teriflunomide | 2 |
| Cladribine | 1 |
| Ocrelizumab | 1 |
| No therapy | 5 |

*Note:* Abbreviations: EDSS, Expanded Disability Status Scale; 9HPT, Nine Hole Peg Test; TMT-A, Trail Making Test part A; TMT-B, Trail Making Test part B; SD, standard deviation.

### MRI

Scanning was performed in a 3T Siemens TrioTim MRI scanner with a 32-channel head coil (Siemens Healthcare, Erlangen, Germany). A 3D T1-weighted magnetization prepared rapid gradient echo (MP-RAGE) sequence (echo time [TE] = 2.52 ms, repetition time [TR] = 1900 ms, inversion time [TI] = 900 ms, flip angle = 9°, matrix size = 256 × 256, field of view [FOV] = 256 × 256 mm^2^, slice thickness = 1 mm, voxel size = 1 × 1 x 1 mm^3^) and a sagittal 3D turbo spin-echo fluid attenuated inversion recovery (FLAIR) sequence (TE = 388 ms, TR = 5000 ms, TI = 1800 ms, flip angle = 120°, matrix size = 256 × 258, FOV = 256 × 256 mm^2^, slice thickness = 1 mm, voxel size = 0.5 × 0.5 × 1 mm^3^) were acquired. Further, multi-shell DWI data were obtained (TE = 111 ms, TR = 10800 ms, flip angle = 90°, matrix size = 128 × 128, FOV = 2304 × 2304 mm^2^, slice thickness = 2 mm, voxel size = 2 × 2 x 2 mm^3^), with three non-zero *b*-values each measured in thirty unique directions (*b* = 900 s/mm^2^, *b* = 1800 s/mm^2^, *b* = 2700 s/mm^2^) in the anterior-posterior direction. Six non diffusion-weighted volumes were acquired before each change in *b*-value in the anterior-posterior direction. Finally, one further non diffusion-weighted volume was acquired in anterior-posterior, and one in posterior-anterior direction.

### Transcranial magnetic stimulation and recordings of motor evoked potentials

Electromyography (EMG) electrodes were placed over the right first dorsal interosseous (FDI) and abductor pollicis brevis (APB). EMG signals including the motor evoked potentials (MEP) were 1000-fold amplified with a D440 amplifier from Digitimer (Fort Lauderdale, USA). Using a CED 1401 laboratory interface, signals were digitized with a sampling rate of 5 kHz (Cambridge Electronic Design, Cambridge, UK). TMS was applied using a Rapid^2^ Stimulator with a figure-eight coil in biphasic pulse configuration (Magstim®, Whitland, UK). The TMS coil was placed over the hand area of left primary motor cortex (M1) approximately in parallel to the central sulcus, i.e. 45-55° relative to the mid-sagittal line (Groppa et al., 2012). After each suprathreshold pulse consistently evoked an MEP in the right FDI and APB, the individual resting motor threshold was determined. To that end, pulse intensity was lowered until MEPs with peak-to-peak amplitudes of 50 *μV* were evoked for two out of four pulses at rest (Wilson et al., 1993).

### Neuropsychological assessment

In the MS group, 31 patients underwent neuropsychological cognitive assessment of 9HPT, and 44 patients of TMT-A and TMT-B (time interval between MRI and neuropsychological testing was mean = 3.8 months, SD = 3.1 months). The 9HPT is a frequently used measure of manual dexterity that is often impaired in MS (Feys et al., 2017). Participants are asked to place and remove pegs into holes on a board as quickly as possible. We used 9HPT scores of the dominant (right) hand as we accordingly investigated the left-hemispheric M1 cortex. In the TMT, patients are instructed to connect a set of numbers and letters according to specific rules as fast as possible. Whereas we included the test due to the evaluation of motor speed, it also requires visual attention and task switching abilities. Average raw scores and standard deviations are reported in Table 1. We used z-scores of these variables by comparing the raw scores with test-specific normative data stratified for age and education.

### Data preprocessing

Figure 1 provides an overview of the study pipeline.

**Figure 1.**
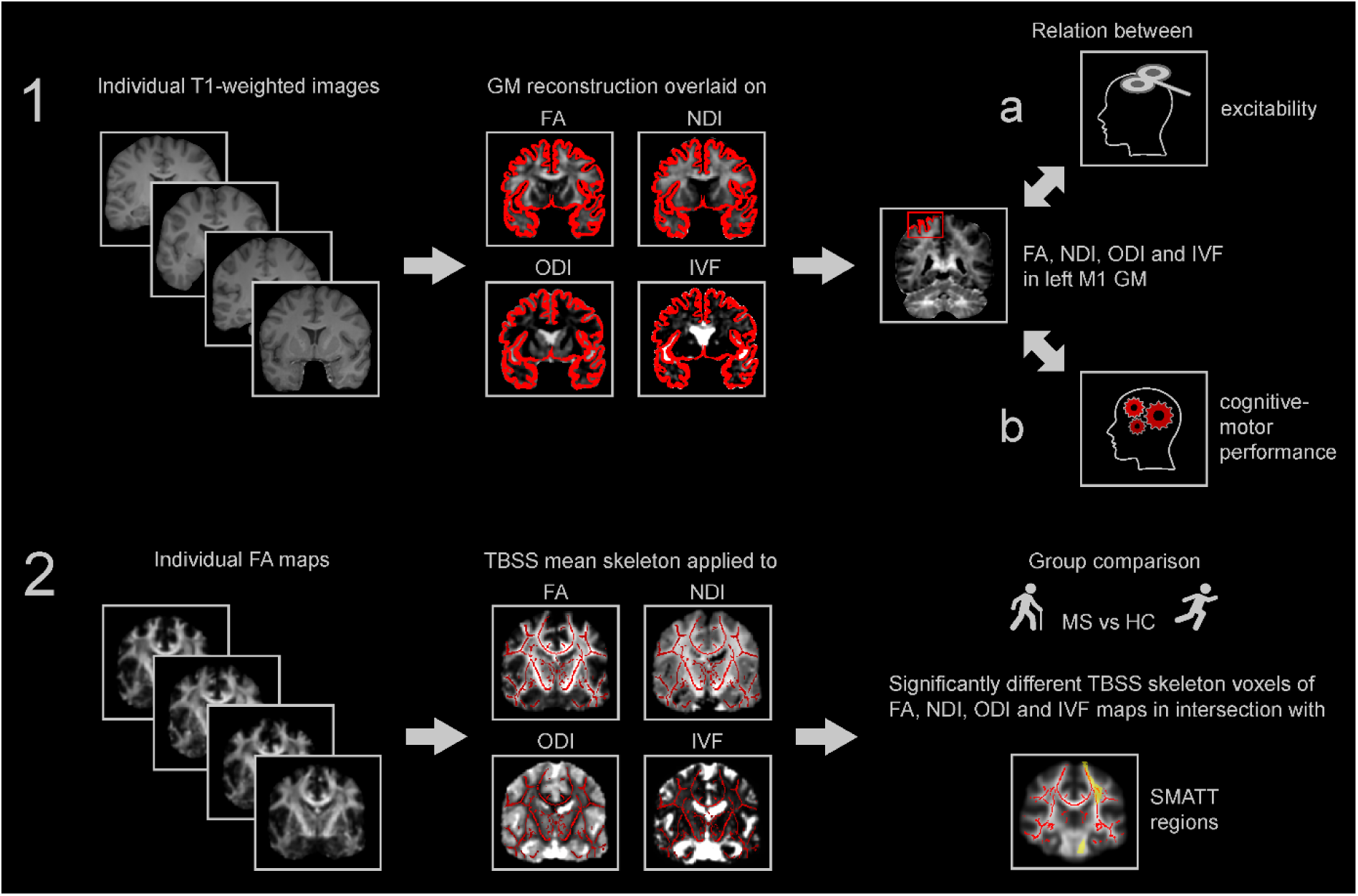
Study pipeline. (1) T1-weighted images were used for reconstruction of the GM that was individually coregistered to FA, NDI, ODI and IVF maps. Motor threshold as measure of excitability was regressed on average FA, NDI, ODI and IVF within the GM of left M1 (1a). Scores of cognitive-motor performance were similarly regressed on average FA, NDI, ODI and IVF within the GM of left M1 (1b). (2) Individual FA maps of all subjects were used to compute the TBSS mean skeleton that was applied to FA, NDI, ODI and IVF for comparisons of MS and HC group. Percentage of intersecting significantly different voxels with SMATT regions were determined for each diffusion parameter. Abbreviations: FA, fractional anisotropy, NDI, neurite density index, ODI, orientation dispersion index, IVF, isotropic volume fraction, MS, multiple sclerosis, HC, healthy control, TBSS, tract-based spatial statistics, SMATT, sensorimotor area tract template, GM, grey matter, M1, primary motor cortex.

### Brain segmentation

Brain segmentation based on T1-weighted images was conducted using the cross-sectional processing stream of FreeSurfer version 5.3.0 (http://surfer.nmr.mgh.harvard.edu/). Here, removal of non-brain tissue, automated Talairach transformation, cortical and subcortical segmentation, intensity normalization, tessellation of the grey and white matter boundary and automated topology correction is included (Fischl, 2012). We used the FreeSurfer-based GM segmentation for further analyses.

### Preprocessing of DWI

DWI data were preprocessed using the diffusion toolbox of FSL version 5.0.9 (https://fsl.fmrib.ox.ac.uk/fsl/) by correcting susceptibility induced distortions using *topup* and applying eddy current and motion artifact correction using *eddy*. Voxel-wise diffusion tensor fitting and computation of FA was performed using FSL’s *dtifit* only considering the inner shell (*b* = 900) and the non diffusion-weighted images.

### Application of the NODDI model

Zhang and colleagoues developed the NODDI model with the requirement of being sufficiently simple, while being complex enough for depiction of major characteristics of neurite morphology (Zhang et al., 2012). A further aim was a clinically feasible acquisition time below 30 minutes. We fitted the model to the DWI using the Accelerated Microstructure Imaging via Convex Optimization (AMICO) algorithm resulting in NDI, ODI and IVF maps (Daducci et al., 2015). Whereas IVF is characterized by free isotropic Gaussian diffusion, NDI and ODI are computed from restricted diffusion adapted from the orientation-dispersed cylinder model (Zhang et al., 2011, 2012). We coregistered the first non diffusion-weighted image to the brainmask obtained with FreeSurfer using FSL’s FMRIB’s Linear Image Registration Tool (*FLIRT*). The transformation matrix was then applied to FA, NDI, ODI and IVF maps.

### Application of a motor atlas

The Human Motor Area Template (HMAT) contains six sensorimotor regions within each hemisphere (Mayka et al., 2006). As the left M1 mask of the HMAT includes both GM and WM, we first also coregistered it to the brainmask in FreeSurfer space. This allowed us to compute average values of FA, NDI, ODI and IVF exclusively within the GM of left M1 as reconstructed in FreeSurfer.

### Lesion segmentation

We estimated lesion volumes with the lesion growth algorithm (Schmidt et al., 2012) implemented in the lesion segmentation toolbox (LST) version 2.0.15 (https://www.applied-statistics.de/lst.html). T2-FLAIR volumes were coregistered to the T1-weighted images and segmentation information used to compute lesion belief maps. After visual inspection, κ = 0.2 was chosen for thresholding of these maps. By growing lesions along hypointensely appearing voxels in the T2-flair images, binary lesion maps were obtained. We linearly transformed T2-flair images to the brainmask obtained with FreeSurfer using FSL’s *FLIRT*. The transformation matrix was applied to transform lesion maps to FreeSurfer space. The percentage of lesion load in the left M1 mask was computed and used as covariate for regression analyses.

### Statistical analyses

#### 1. Microstructural correlates of motor excitability and cognitive-motor performance

Normality was assessed visually and using the Lilliefors test in *R* (version 3.6.3, *RStudio*). For group comparisons of motor threshold, FA, NDI, ODI and IVF, t-tests were conducted and considered significant in case of *p* < .05. Additionally, we tested homogeneity of variance between the groups using Levene’s test. Correlation coefficients were computed in *R* using Pearson correlation. We used *R* package *corrplot* for depiction of correlation coefficients (Figure 2 and S1). Hierarchical backwards regression was conducted using *IBM SPSS Statistics for Windows*, Version 23 (IBM Corp., Armonk, NY) for the MS and HC group separately. As control variables, we entered gender and age in the model for prediction of the dependent variable motor threshold. Additionally, the percentage of lesion load in left M1 was considered as control variable for the MS model. All control variables were entered first in one block in the model. Variables of interest were FA, NDI, ODI and IVF averaged within the left M1 GM mask. Similarly, regression with backwards elimination was conducted for 9HPT, TMT-A and TMT-B scores on FA, NDI, ODI and IVF in left M1 GM in MS patients. The order of removal of variables of interest was determined based on increase in correlation coefficients with the dependent variable (Figure 2 and S1), and predictors were only retained in case of *p* < .05 of the regression coefficient.

**Figure 2.**
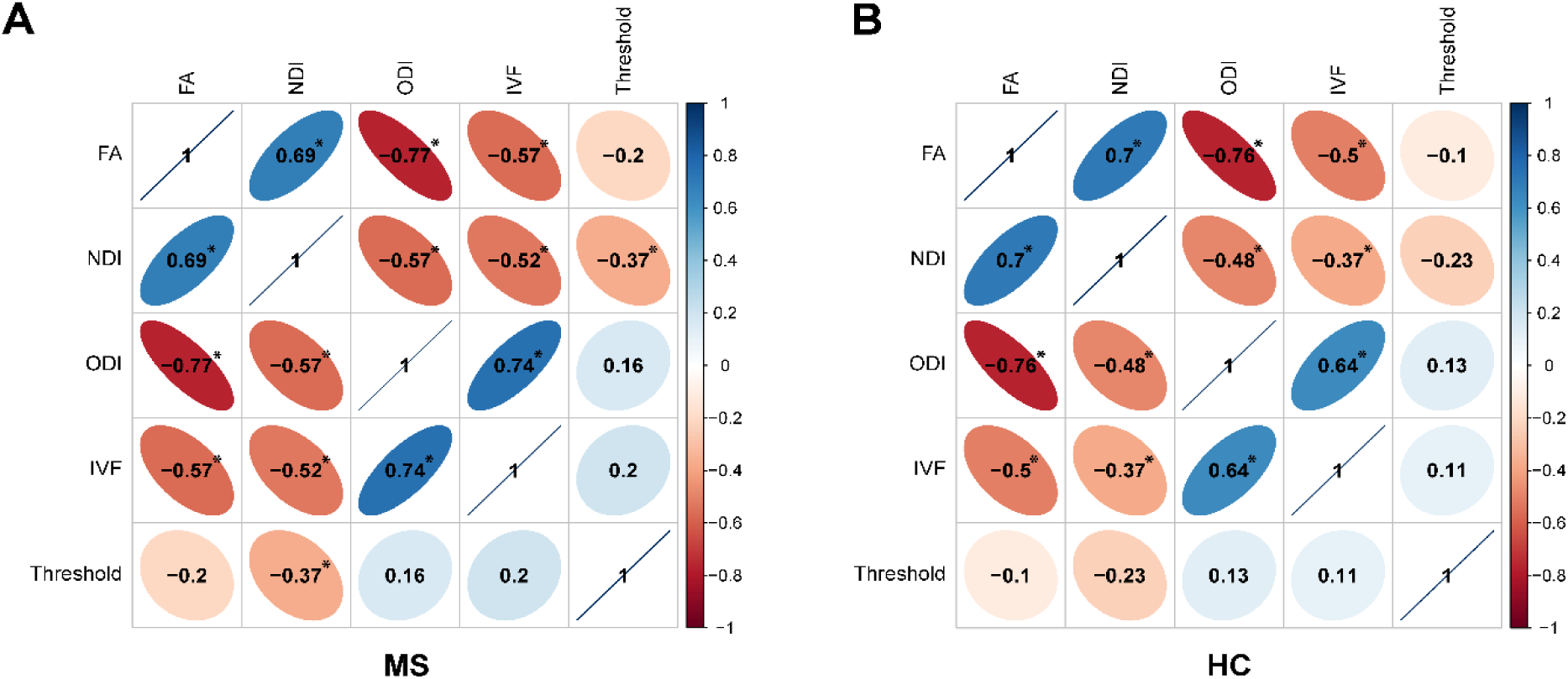
Correlation coefficients of the correlation between left M1 GM FA, NDI, ODI, IVF and motor threshold in (A) MS and (B) HC. * *p* < .05. Abbreviations: M1, primary motor cortex, GM, grey matter, FA, fractional anisotropy, NDI, neurite density index, ODI, orientation dispersion index, IVF, isotropic volume fraction, MS, multiple sclerosis, HC, healthy controls.

#### 2. White matter microstructural integrity of the motor system

First, TBSS was conducted with FA maps of both groups. This method allows voxel-wise group comparisons by non-linear registration and projection onto an alignment-invariant tract representation (Smith et al., 2006). As target for the nonlinear registration, the FMRIB58_FA_1mm standard space template as provided by FSL was used. The skeleton was thresholded to κ = 0.2 (Smith et al., 2006). The resulting skeleton was then also applied to NDI, ODI and IVF maps. Permutation test using the randomize tool was applied for statistical analyses (Winkler et al., 2014). For each group comparison of FA, NDI, ODI and IVF between HC and MS, 500 permutations were carried out with the threshold-free cluster enhancement (TFCE) option to control for multiple compairsons. A threshold of *p* < .05 was used to define significance (Smith & Nichols, 2009). We overlaid all tract masks of the sensorimotor area tract template (SMATT) (Archer et al., 2018) on the mean FA skeleton and computed the number of voxels in the skeleton for each SMATT region. For creation of the SMATT, HMAT regions were used as seed region with waypoints placed at the level of the posterior limb of the internal capsule and the cerebral peduncle under exclusion of transcallosal fibers (Archer et al., 2018). We then extracted the number and percentage of voxels with significant group differences intersecting with SMATT tracts for FA, NDI, ODI and IVF maps, respectively. Lastly, we computed the percentage of overlapping group differences in all diffusion parameters per region and averages over all regions.

In Supplementary Results S5, we additionally provide results of a congruent analysis regarding Johns Hopkins University (JHU) DTI-based WM atlas that also covers tracts outside the motor system (Hua et al., 2008; Mori et al., 2005; Wakana et al., 2007).

## Results

HC and MS were all right-handed and did not significantly differ in age or gender distribution (both *p* > .05).

### 1a. Microstructural correlates of motor cortex excitability

We first investigated group differences of motor threshold and FA, NDI, ODI and IVF in the left M1 GM between the MS and HC groups. Excitability levels, i.e. motor thresholds were non-significantly higher in MS patients compared to HC (t(97) = 0.78, *p* > .05; MS group: mean [SD] = 58.06 [10.60]; HC group: mean [SD] = 56.55 [8.63]). Group comparisons in FA, NDI, ODI and IVF in the GM of left M1 all did not reveal any significant group differences (all *p* > .05). Variability of NDI in MS patients was significantly higher than in HC (F(1,97) = 4.25, p = .04).

Within the GM of both MS and HC, diffusion measures were significantly interrelated (Figure 2). FA and NDI were positively interrelated, but negatively correlated to ODI and IVF. In MS patients, higher NDI was significantly linked to lower motor threshold; HC exhibited a similar trend.

We first entered age and gender as potentially confounding variables into the hierarchical backward regression model for the prediction of motor threshold for MS and HC separately. Predicting motor threshold in the MS group, we additionally included lesion load in M1. As variables of interest, we considered FA, NDI, ODI and IVF in the GM of left M1, which served as the stimulation site for determination of the motor threshold. NDI in the GM of left M1 significantly predicted motor threshold in MS (F(1,48) = 7.493, *p* = .009) and explained 11.7% of its variance (for details of hierarchical regression analysis see Table S2). Figure 3A depicts the relationship between the two variables and the respective density estimates, showing that lower neurite density in MS is linked to higher motor threshold, i.e. lower excitability. No model was significant in the HC group, however there was similarly a trend for NDI predicting motor threshold (Figure 3B, Table S3). By including NDI of both groups and a dummy variable encoding group membership for the prediction of motor threshold, we tested significance of group-specific slopes. The coefficient of the interaction term was not significant (*p* > .05), indicating that slopes were not significantly different between MS patients and HC.

**Figure 3.**
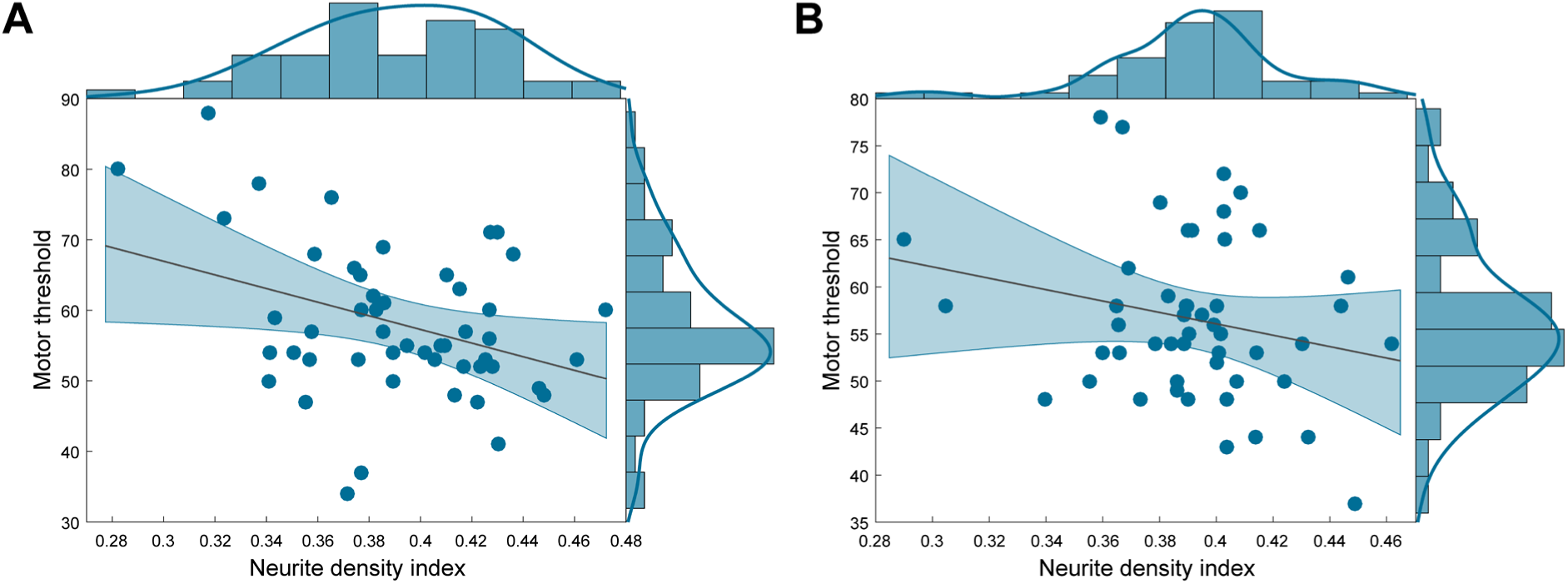
Scatterplot with regression line and density estimates of the regression of motor threshold on neurite density index in (A) patients with MS and (B) HC. Abbreviations: HC, healthy controls, MS, multiple sclerosis.

### 1b. Microstructural correlates of cognitive-motor performance

MS patient’s 9HPT scores were regressed on control variables age, gender and lesion load in M1 and variables of interest FA, NDI, ODI and IVF in left M1 GM. No variable of interest significantly predicted 9HPT scores (Table S4).

Equivalently, TMT z-scores were regressed on control variables age, gender and lesion load in M1 and variables of interest FA, NDI, ODI and IVF in left M1 GM. NDI in the GM of left M1 was a significant predictor of both TMT-A (F(1,42) = 8.102; *p* =.007; adjusted r^2^ = .142) and TMT-B z-scores (F(1,42) = 7.390; *p* =.009; adjusted r^2^ = .129) (for details of hierarchical regression analyses see Table S5 and S6). Lower NDI predicted worse performance in both TMT-A (Figure 4A) and TMT-B (Figure 4B).

**Figure 4.**
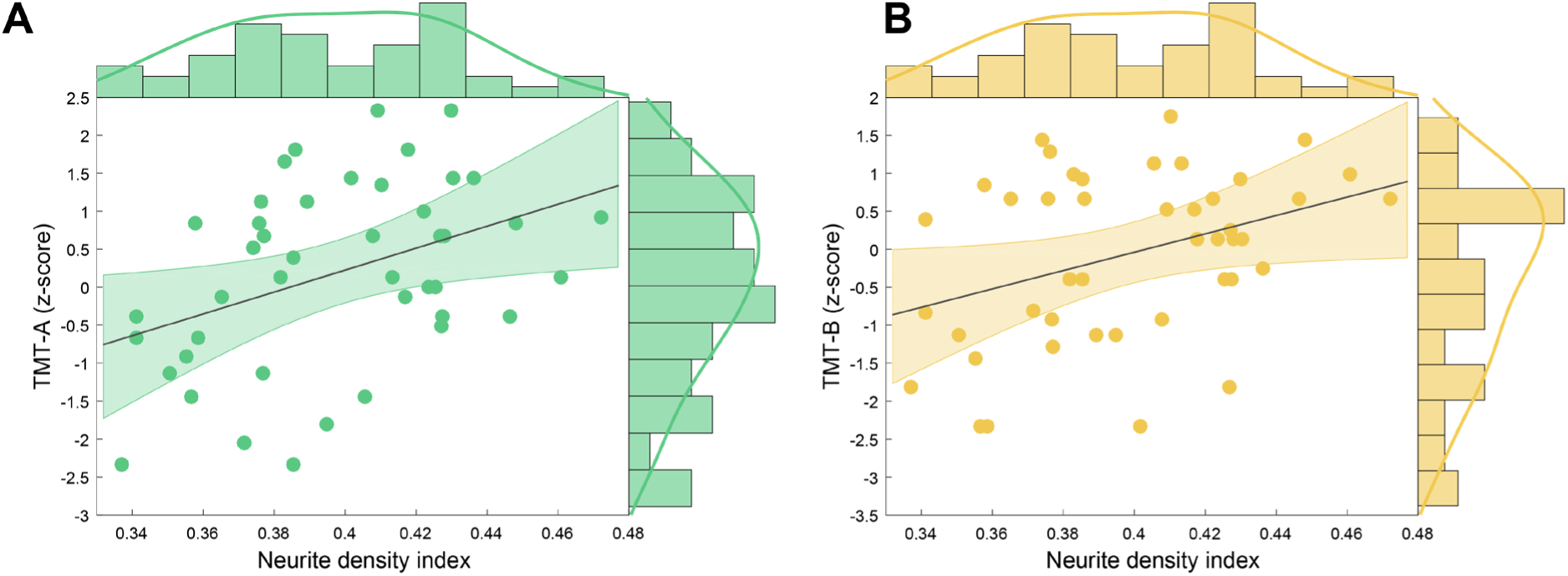
Scatterplot with regression line and density estimates of the regression of (A) TMT-A (z-scores) and (B) TMT-B (z-scores) on neurite density index in patients with MS. Abbreviations: MS, multiple sclerosis, TMT-A, Trail Making Test part A, TMT-B, Trail Making Test part B.

### 2. White matter microstructural integrity of the motor system

First, we compared FA, NDI, ODI and IVF maps between HC and MS patients using TBSS. TFCE-based significance tests showed significantly higher FA and NDI values in HC compared to MS in a large amount of motor WM tract voxels, i.e. 6.2% and 10.9% of the mean motor skeleton, respectively (*p* < .05). Conversely, ODI was higher in MS patients compared to HC in 3% of the mean motor skeleton voxels (*p* < .05) (Figure 5). No voxels showed significantly increased NDI or decreased ODI in MS compared to HC (*p* > .05). No significant group differences were observed regarding IVF (*p* > .05).

**Figure 5.**
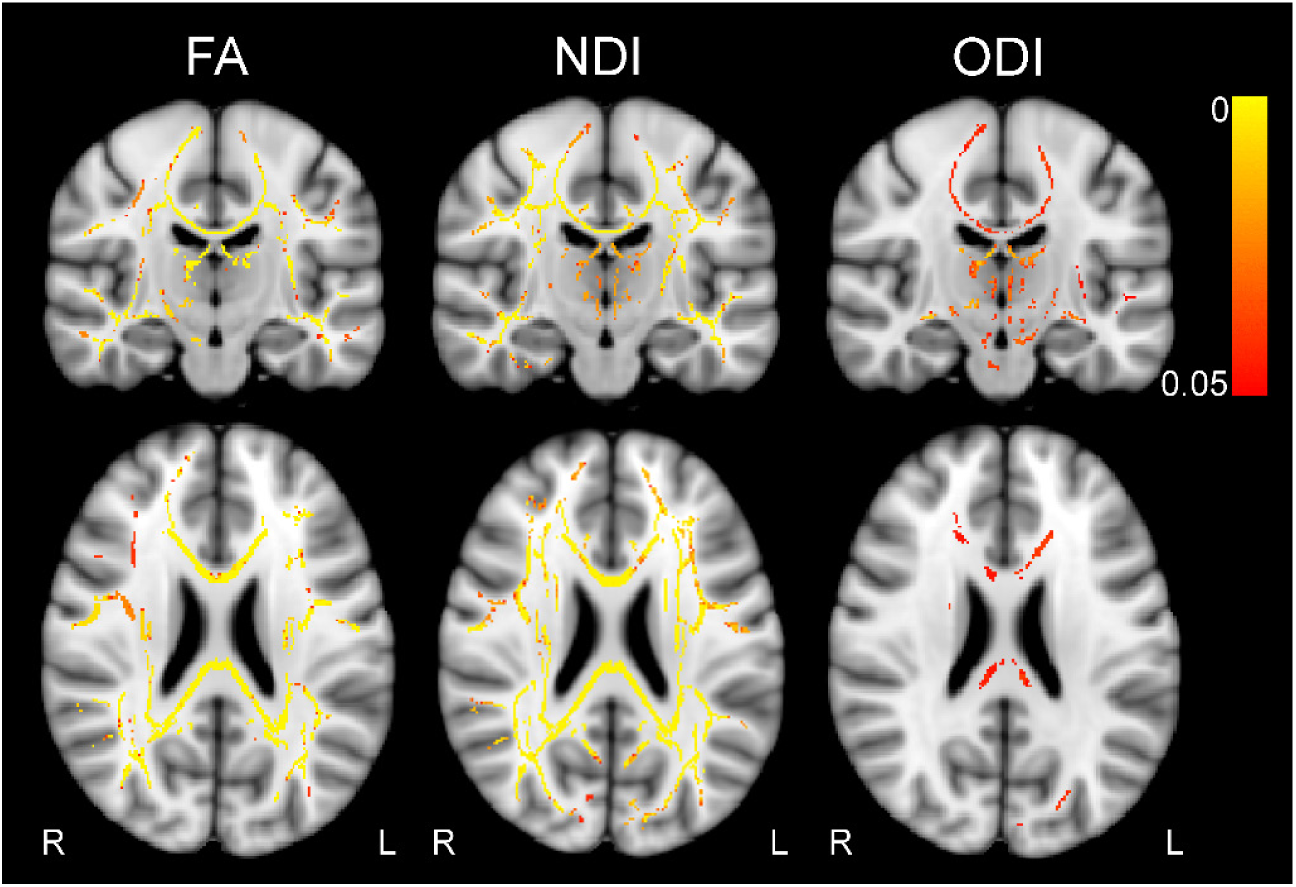
Results of tract-based spatial statistics analyses. Color bar indicates TFCE-corrected *p*-value, thresholded at *p* < .05. Contrast HC > MS for FA and NDI, and MS > HC for ODI. MNI coordinates: x = 90, y = 109, z = 95. Abbreviations: FA, fractional anisotropy, NDI, neurite density index, ODI, orientation dispersion index, L, left, R, right, TFCE, threshold-free cluster enhancement.

To obtain more clarity regarding the regional effects, we computed the number and percentage of voxels showing group differences for each region of the SMATT atlas for FA, NDI and ODI maps (Table 2). Fibers exhibiting group differences in at least 10% of the region in either FA, NDI or ODI included right supplementary motor area (SMA), pre-SMA, ventral premotor cortex (PMv) and bilateral somatosensory cortex (S1) and M1.

**Table 2.** Number and percentage of voxels significantly differing between MS and HC in FA, NDI and ODI in SMATT regions. The last column contains the total number of voxels of the mean skeleton intersecting with each SMATT region. Contrast HC > MS for FA and NDI, and MS > HC for ODI, sorted decreasingly by the percentage of voxels showing significant group differences of each region, summed over FA, NDI and ODI

| **SMATT label** | **FA (%)** | **NDI (%)** | **ODI (%)** | **Total** |
| --- | --- | --- | --- | --- |
| Right SMA | 571 (11.28) | 605 (11.95) | 308 (6.09) | 5061 |
| Right M1 | 706 (8.17) | 1348 (15.59) | 377 (4.36) | 8644 |
| Right S1 | 363 (9.96) | 391 (10.73) | 198 (5.44) | 3643 |
| Left M1 | 481 (5.56) | 1275 (14.75) | 121 (1.40) | 8644 |
| Right PMv | 379 (10.02) | 350 (9.26) | 84 (2.22) | 3781 |
| Right PMd | 296 (7.00) | 342 (8.09) | 215 (5.08) | 4230 |
| Right pre-SMA | 283 (4.95) | 669 (11.70) | 104 (1.82) | 5720 |
| Left SMA | 240 (4.74) | 410 (8.10) | 263 (5.20) | 5061 |
| Left S1 | 182 (3.18) | 681 (11.91) | 47 (0.82) | 5720 |
| Left pre-SMA | 143 (3.93) | 287 (7.88) | 140 (3.84) | 3643 |
| Left PMv | 223 (5.90) | 308 (8.15) | 48 (1.27) | 3781 |
| Left PMd | 10 (0.24) | 80 (1.89) | 101 (2.39) | 4230 |
| **TOTAL** | **3877 (6.24)** | **6746 (10.85)** | **2006 (3.09)** | **62158** |

*Note:* Abbreviations: MS, multiple sclerosis, HC, healthy control, FA, fractional anisotropy, NDI, neurite density index, ODI, orientation dispersion index, M1, primary motor cortex, SMA, supplementary motor area, S1, primary somatosensory cortex, PMd, dorsal premotor cortex, PMv, ventral premotor cortex.

Next, we disentangled which combination of FA, NDI and ODI, or if one of the parameters alone contributed to observed group differences. Figure 6A depicts the percentage of all combinations and single parameters for each SMATT region, while the percentage averaged over all regions is presented in Figure 6B. Neurite density exclusively (56%) and in combination with FA (19%) accounted for the largest amount of differences, followed by ODI alone (9%).

**Figure 6.**
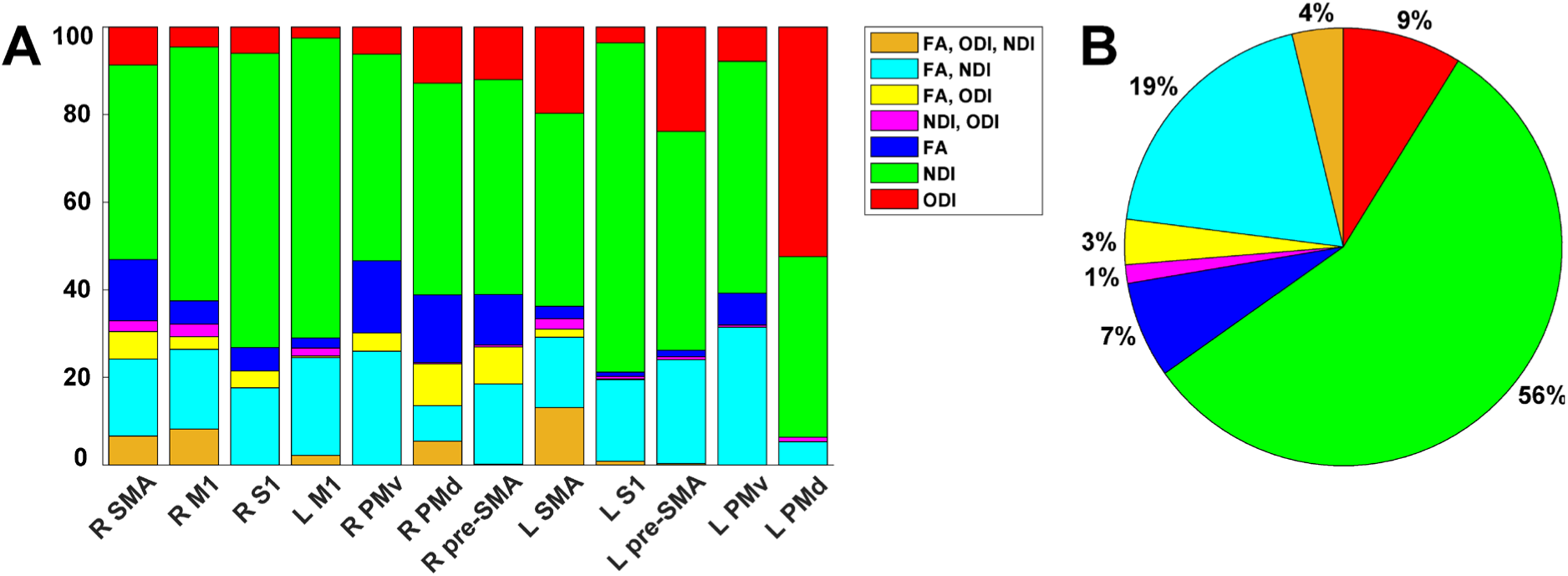
Comparison of diffusion parameters contributing to observed group differences. (A) Percentage of FA, NDI and ODI separately or combined showing group differences in the tract skeleton in intersection with SMATT regions. (B) Percentage averaged over regions with any significant group difference (B). Abbreviations: FA, fractional anisotropy, NDI, neurite density index, ODI, orientation dispersion index, SMA, supplementary motor area, M1, primary motor cortex, S1, primary somatosensory cortex, PMv, ventral premotor cortex. PMd, dorsal premotor cortex.

## Discussion

In order to link microstructural integrity and cortical excitability, we applied a biophysical diffusion model on the basis of a multi-shell acquisition protocol. This allowed us to obtain maps of NDI, ODI and IVF in addition to the classical DTI parameter FA within the WM and GM of the brain motor regions. We linked lower microstructural integrity in the GM to lower motor cortical excitability and lower cognitive-motor performance in MS. In the WM, we observed lower FA and NDI and higher ODI in MS patients compared to HC in all motor tracts. Voxel-wise group differences in FA strongly co-occurred with NDI differences. NDI and ODI appear sensitive for detection of neuropathological changes in MS patients undetected by FA.

### 1a. Microstructural correlates of motor excitability

Cortical excitability can be altered in MS patients following neuronal and axonal loss (Calabrese et al., 2007; Zipser et al., 2018). To unravel the link between microstructure and excitability, we applied NODDI, a biophysical model of microstructure. We postulate a tight excitability-microstructural integrity relationship that strengthens during the disease course. A robust quantification of this relationship could predict functional impairment and be extended to monitor the disease course or therapeutic responses.

In contrast to classical DTI-based measures such as FA that are biased in areas of pronounced neurite orientation dispersion such as the GM, this tendency is mitigated in the NODDI model that is inherently more specific in depicting the cortical microstructure (Zhang et al., 2012). We were interested in how motor cortical integrity quantified by NODDI parameters explains altered excitability as evaluated by the resting motor threshold, a unique non-invasive approach cortex excitability assessments in humans. An initial comparison between the MS and HC groups revealed that motor threshold exhibited a trending increase in MS compared to HC. Higher motor thresholds, i.e. lower excitability, in MS than HC have been reported previously, particularly for the secondary progressive disease type and in patients in the relapsing phase (Caramia et al., 2004; Vucic et al., 2012). Decreased motor cortical excitability in MS patients potentially is a consequence of neuronal loss, axonal scarcity or synaptic down-scaling of the cortico-cortical inputs (Boorman et al., 2007; Calabrese et al., 2007; Wahl et al., 2007; Zipser et al., 2018). The motor thresholds did not statistically differ between the addressed groups in our study, however showed a trending increase in MS. All diffusion measures did not significantly depict group differences, but in line with a previous study, trending lower ODI in MS compared to HC was observable (De Santis et al., 2019). A decrease in the GM ODI could indicate degeneration of single neurites, resulting in dispersion reductions (Spano et al., 2018; Zhang et al., 2012). Presumably, both a larger sample size and a longer disease duration would amplify these trending differences in motor threshold and NODDI parameters between MS and HC.

We next examined the relation of FA, NDI, ODI and IVF among each other within M1, where the stimulation for assessment of the motor threshold was conducted. FA and NDI were significantly positively interrelated, and negatively linked to ODI and IVF. A hierarchical backward regression model revealed that lower NDI predicted higher motor threshold, i.e. lower neural excitability in MS patients. This is well in line with the suggestion of axonal and myelin loss due to neuroinflammatory processes (Filippi et al., 2012; Grussu et al., 2017). Furthermore, this shows the superiority of NDI in predicting excitability over local lesion load that was included in the model as a potential confounder. The HC model was not significant, however a trend of a positive relation between NDI and motor threshold was observable here as well. We suggest that higher scattering of NDI led to a better model fit in the MS group as higher variance allows stronger correlations, while the model in general also fits to the HC group. This assumption could be tested by involving HC with more strongly scattered values, such as by including individuals of higher age that typically show higher resting motor thresholds and increasing neuronal loss (Bhandari et al., 2016; Ding et al., 2016; Merluzzi et al., 2016).

### 1b. Microstructural correlates of cognitive-motor performance

We further assessed if microstructural integrity of the left M1 GM can also be linked to a behavioral outcome, i.e. cognitive-motor performance in the MS group. Hierarchical regression results indicated that patients with a loss in neurite density showed a lower cognitive-motor performance as quantified by TMT-A and TMT-B. For 9HPT, the regression model did not include significant predictors of interest. However, fewer patients completed this test and a trend for a positive correlation was detected between NDI and 9HPT scores (Figure S1). Previous results demonstrated a relation between lower cognitive performance and lower GM volume in regions relevant for demands of the utilized neuropsychological test (Morgen et al., 2006), but also the whole brain (Fisniku et al., 2008; Sastre-Garriga et al., 2004). Lower magnetization transfer ratio was observed in motor regions for patients with lower 9HPT performance, likely reflecting pathological processes including inflammation, edema, demyelination and axonal loss (Khaleeli et al., 2007; Schmierer et al., 2004; van Waesberghe et al., 1999). The TMT measures multiple cognitive domains such as visuo-motor abilities and cognitive flexibility and it has been shown that precentral gyrus was activated in an fMRI-compatible TMT adaptation (Crowe, 1998; Zakzanis et al., 2005). Hence, a lowered TMT performance linked to lower NDI in left M1 as seen here appears consequential and underlines the suitability of the NODDI model for assessment of GM microstructural integrity.

### 2. White matter microstructural integrity of the motor system

To gain a complete depiction of motor-relevant microstructure and its pathological alterations in MS, we next investigated WM motor tracts by applying TBSS. We detected decreased FA and NDI and increased ODI in patients with MS compared to HC, where the highest percentage of voxels showing group differences was in NDI. The strongest portion of sensorimotor tracts in intersection with group differences in the TBSS FA mean skeleton included WM tracts originating in right SMA and PMv. NDI was particularly lower in bilateral M1 tracts, whereas ODI was higher mainly in right SMA and S1 tracts of MS patients. GM atrophy in MS compared to HC in precentral and postcentral regions has been reported previously (Lansley et al., 2013; Prinster et al., 2010). In a recent longitudinal study, M1 and SMA atrophy was more pronounced in patients with disability progression compared to those without after 10 years (Bergsland et al., 2018). These findings make it plausible that connecting WM tracts show abnormalities due to primary or secondary causes as well, which has been demonstrated earlier (Steenwijk et al., 2015). Also in support of this view, lower FA was observed in the corticospinal tract of MS patients, and higher mean, axial and radial diffusivity was linked to lower M1 thickness (Bergsland et al., 2015). To the best of our knowledge, NODDI has not yet been investigated systematically in sensorimotor tracts of MS patients as was done in the present work. However, a small sample-sized first NODDI study also reported decreased NDI and increased ODI in the normal-appearing WM of MS patients (Schneider et al., 2017). Higher ODI indicates fiber coherence loss and can possibly be explained by an increase in compensative axonal sprouting or branching (Timmers et al., 2015). In a recent histological study, ODI was shown to indeed reflect the neurite architecture complexity, whereas reduced NDI indicates loss of myelin and axons (Grussu et al., 2017). Regarding NDI, a decrease in MS is expected due to the demyelinating, inflammatory and neuro-axonal pathology of the disease (Filippi et al., 2012). A NODDI study that performed voxel-wise comparisons revealed reduced NDI and increased ODI in the fornix and corpus callosum of RRMS compared to HC (Spano et al., 2018). Our supplementary results applying the same analysis to JHU regions similarly yielded reduced NDI and increased ODI in MS compared to HC in these structures.

Importantly, low FA values could be caused by lower NDI, higher ODI, or a combination of both (Zhang et al., 2012), such that it is of great relevance to disentangle if group effects appear for these parameters exclusively in a voxel, or if they potentially overlap, which has not yet been systematically quantified in MS research to our knowledge. This allows a more concrete inference on the type of microstructure change in contrast to assumptions solely based on FA. On average, 56 % of voxels showing significant group differences in intersection with the mean FA skeleton were different exclusively in NDI, whereas 19 % were different simultaneously in FA and NDI, 9 % in ODI only and 7 % in FA only. This pattern was consistent among single sensorimotor WM tracts. Our findings suggest that FA is capable of detecting parts of the pathological alterations that can mainly be retraced to NDI decreases in MS. The NODDI model therefore allows to capture further microstructure changes in MS in addition to its inherently higher specificity to the underlying pathology (Zhang et al., 2012). These TBSS results support previous findings based on voxel-wise comparisons (Schneider et al., 2017; Spano et al., 2018) and further disentangle that the lower FA signal is predominantly caused by reduced neurite density in sensorimotor WM tracts of MS patients.

### Limitations and prospect

We provide novel insights regarding the applicability of the NODDI model to assess microstructure in both GM and WM. Our suggestion that neurite density is also linked to motor cortical excitability in healthy participants needs to be further examined in future studies including a subject group with a higher variability in age. Furthermore, not all MS patients completed the 9HPT, which likely lowered statistical power. The link between neuropsychological test performance and NODDI parameters could in future be expanded to other cognitive domains and brain regions analyzing larger patient samples. It would be particularly interesting to observe potential effects of motor and cognitive training or medication on beneficial adaptations in microstructural integrity (Bonzano et al., 2014; Moccia et al., 2017). Due to the multiple facets of the disorder, including the specific disease type and duration, treatment, relapse rate and localization of pathologically affected regions, investigating larger sample sizes that allow stratification to control these variations is of importance. How microstructural state as evaluated by biophysical diffusion models is connected to functional adaptive and maladaptive mechanisms of brain networks would be an interesting question to revisit using functional MRI (Droby et al., 2016; Fleischer et al., 2020; Fleischer, Radetz, et al., 2019). Moreover, the integration of further structural imaging methods such as relaxation times and other diffusion models is relevant for in vivo delineation of pathological microstructure changes as are histological assessments in ex vivo samples (Alexander et al., 2019; Gracien et al., 2017; Grussu et al., 2017; Lakhani et al., 2020). A translational approach linking NODDI in different animal models of MS to human patients remains to be further pursued as was done with a cuprizone model of demyelination in mice and by applying DTI (Cerina et al., 2020; Krämer et al., 2019; Wang et al., 2019). Furthermore, differentiating the evolution of microstructure in MS compared to healthily aging individuals is of interest in longitudinal investigations (Fleischer, Koirala, et al., 2019).

### Conclusion

In this work we applied a biophysical model of GM and WM microstructure in the motor system of patients with MS and age-matched HC. We hereby demonstrate that lower neurite density in left M1 is linked to decreased motor cortical excitability in patients with MS. Also on the behavioral level, decreased neurite density is indicative of impaired cognitive-motor performance. Furthermore, we showed that lower neurite density and higher orientation dispersion are characteristic in the WM of MS patients compared to HC, and that these markers are more sensitive to pathological alterations than the classical DTI measure FA. These findings suggest that advanced biophysical diffusion models are of great relevance for prediction of neurodegenerative processes, excitability alterations and disease progression.

## Supporting information

Supplementary Results

## Data Availability

The data that support the findings of this study are available from the corresponding author upon reasonable request.

## Abbreviations

APB: Abductor pollicis brevis
AMICO: Accelerated Microstructure Imaging via Convex Optimization
DTI: diffusion tensor imaging
DWI: diffusion-weighted imaging
PMd: dorsal premotor cortex
TE: echo time
EMG: electromyography
EDSS: Expanded Disability Status Scale
FOV: field of view
FDI: first dorsal interosseous
FLAIR: fluid attenuated inversion recovery
FA: fractional anisotropy
GM: grey matter
HC: healthy controls
HMAT: Human Motor Area Template
TI: inversion time
IVF: isotropic volume fraction
JHU: Johns Hopkins University
L: left
LST: lesion segmentation toolbox
MEP: motor evoked potential
MS: multiple sclerosis
NDI: neurite density index
NODDI: neurite orientation dispersion and density imaging
9HPT: Nine Hole Peg Test
ODI: orientation dispersion index
M1: primary motor cortex
S1: primary somatosensory cortex
TR: repetition time
RRMS: relapsing-remitting MS
R: right
SMATT: sensorimotor area tract template
SD: standard deviation
SMA: supplementary motor area
TFCE: threshold-free cluster enhancement
TBSS: tract-based spatial statistics
TMT-A: Trail Making Test part A
TMT-B: Trail Making Test part B
TMS: transcranial magnetic stimulation
PMv: ventral premotor cortex
WM: white matter

## Acknowledgements

We thank all participants who took part in the study and Cheryl Ernest for proofreading the manuscript. This study was supported by the German Research Council (DFG, CRC TR-128 to S.B., S.G.M., S.G.). Parts of this research were conducted using the supercomputer Mogon and advisory services offered by Johannes Gutenberg University Mainz (hpc.uni-mainz.de), which is a member of the AHRP and the Gauss Alliance e.V. We gratefully acknowledge the computing time granted on Mogon.

