## Supplementary Results for "Linking microstructural integrity and motor cortex excitability in multiple sclerosis"

*S1. Group differences in FA, NDI, ODI and IVF in left M1 GM*

Table S1. FA, NDI, ODI and IVF in left M1 GM in MS compared to HC

| Diffusion parameter | MS (mean [SD]) | HC (mean [SD]) | t-test | Levene's test |
|---|---|---|---|---|
| **FA** | m = 0.1539 [0.0204] | m = 0.1515 [0.098] | t(97) = -0.61; p > .05 | F(1,97) = 0.14, p > .05 |
| **NDI** | m = 0.3915 [0.0393] | m = 0.3916 [0.0321] | t(97) = 0.01; p > .05 | F(1,97) = 4.25, p = .04 |
| **ODI** | m = 0.5119 [0.0344] | m = 0.5195 [0.0330] | t(97) = 1.11; p > .05 | F(1,97) = $2.3*e^{-4}$, p > .05 |
| **IVF** | m = 0.2889 [0.1112] | m = 0.2820 [0.0825] | t(97) = -0.35; p > .05 | F(1,97) = 2.78, p > .05 |

*Note*: T-test for comparison of group differences in each diffusion parameter, Levene's test for testing of variance inequality. Abbreviations: FA, fractional anisotropy, NDI, neurite density index, ODI, orientation dispersion index, IVF, isotropic volume fraction, M1, primary motor cortex, GM, grey matter, MS, multiple sclerosis, HC, healthy control; SD, standard deviation.

*S2. Detailed results of hierarchical regression analysis of motor threshold in MS*

Table S1. Results of hierarchical regression analysis of motor threshold in MS

| | Regression coefficient | Standard error | Beta coefficient | t statistic | *p* value |
|---|---|---|---|---|---|
| **Model 1** | | | | | |
| Age | .006 | .152 | .007 | .042 | .967 |
| Gender | 4.577 | 3.431 | .212 | 1.334 | .189 |
| M1 lesion load | 4.110 | 3.042 | .209 | 1.351 | .184 |
| ODI | -59.439 | 83.964 | -.193 | -.708 | .483 |
| IVF | 15.148 | 23.349 | .159 | .649 | .520 |
| FA | -35.262 | 134.085 | -.068 | -.263 | .794 |
| NDI | -94.881 | 54.737 | -.352 | -1.733 | .090 |
| | F(7,42) = 1.563; *p* = .173; adjusted $r^2$ = .074 | | | | |
| **Model 2** | | | | | |
| Gender | 4.608 | 3.308 | .213 | 1.393 | .171 |
| M1 lesion load | 4.135 | 2.948 | .210 | 1.403 | .168 |
| ODI | -59.768 | 82.612 | -.195 | -.723 | .473 |
| IVF | 15.564 | 20.860 | .163 | .746 | .460 |
| FA | -35.547 | 132.346 | -.068 | -.269 | .790 |
| NDI | -94.511 | 53.377 | -.350 | -1.771 | .084 |
| | F(6,43) = 1.866; *p* = .109; adjusted $r^2$ = .096 | | | | |

| | | | | | |
|---|---|---|---|---|---|
| **Model 3** | | | | | |
| Gender | 3.647 | 2.958 | .185 | 1.233 | .224 |
| ODI | -48.589 | 83.095 | -.158 | -.585 | .562 |
| IVF | 6.536 | 20.039 | .069 | .326 | .746 |
| FA | -15.899 | 132.991 | -.031 | -.120 | .905 |
| NDI | -95.449 | 53.940 | -.354 | -1.770 | .084 |
| | $F(5,44) = 1.812$; $p = .130$; adjusted $r^2 = .077$ | | | | |
| **Model 4** | | | | | |
| ODI | -25.685 | 81.459 | -.084 | -.315 | .754 |
| IVF | 7.612 | 20.135 | .080 | .378 | .707 |
| FA | 31.527 | 128.041 | .061 | .246 | .807 |
| NDI | -111.908 | 52.564 | -.415 | -2.129 | .039 |
| | $F(4,45) = 1.864$; $p = .133$; adjusted $r^2 = .066$ | | | | |
| **Model 5** | | | | | |
| IVF | 3.939 | 16.262 | .041 | .242 | .810 |
| FA | 54.543 | 104.161 | .105 | .524 | .603 |
| NDI | -112.695 | 51.988 | -.418 | -2.168 | .035 |
| | $F(3,46) = 2.501$; $p = .071$; adjusted $r^2 = .084$ | | | | |
| **Model 6** | | | | | |
| FA | 45.940 | 96.933 | .088 | .474 | .638 |
| NDI | -115.445 | 50.222 | -.428 | -2.299 | .026 |
| | $F(2,47) = 3.798$; $p = .030$; adjusted $r^2 = .103$ | | | | |
| **Model 7** | | | | | |
| NDI | -99.095 | 36.201 | -.367 | -2.737 | .009 |
| | $F(1,48) = 7.493$; $p = .009$; adjusted $r^2 = .117$ | | | | |

*Note*: Abbreviations: MS, multiple sclerosis, M1, primary motor cortex, ODI, orientation dispersion index, IVF, isotropic volume fraction, FA, fractional anisotropy, NDI, neurite density index.

*S3. Detailed results of hierarchical regression analysis of motor threshold in HC*

Table S3. Results of hierarchical regression analysis of motor threshold in HC

| | **Regression coefficient** | **Standard error** | **Beta coefficient** | **t statistic** | ***p* value** |
|---|---|---|---|---|---|
| **Model 1** | | | | | |
| Age | .023 | .157 | .025 | .149 | .882 |

| | | | | | |
|---|---|---|---|---|---|
| Gender | 1.351 | 2.906 | .079 | .465 | .644 |
| FA | 100.837 | 128.311 | .231 | .786 | .436 |
| IVF | 1.212 | 22.554 | .012 | .054 | .957 |
| ODI | 41.867 | 69.387 | .160 | .603 | .549 |
| NDI | -91.277 | 56.875 | -.339 | -1.605 | .116 |
| | F(6,42) = 0.592; *p* = .735; adjusted r² = -.054 | | | | |
| **Model 2** | | | | | |
| Gender | 1.277 | 2.830 | .075 | .451 | .654 |
| FA | 101.521 | 126.762 | .233 | .801 | .428 |
| IVFI | 2.584 | 20.350 | .025 | .127 | .900 |
| ODI | 40.253 | 67.751 | .154 | .594 | .556 |
| NDI | -90.654 | 56.073 | -.337 | -1.617 | .113 |
| | F(5,43) = 0.723; *p* = .610; adjusted r² = -.030 | | | | |
| **Model 3** | | | | | |
| FA | 114.957 | 122.095 | .263 | .942 | .352 |
| IVFI | .846 | 19.800 | .008 | .043 | .966 |
| ODI | 42.934 | 66.876 | .164 | .642 | .524 |
| NDI | -90.221 | 55.555 | -.335 | -1.624 | .112 |
| | F(4,44) = 0.868; *p* = .491; adjusted r² = -.011 | | | | |
| **Model 4** | | | | | |
| IVF | 1.526 | 19.762 | .015 | .077 | .939 |
| ODI | 3.669 | 52.215 | .014 | .070 | .944 |
| NDI | -59.257 | 44.718 | -.220 | -1.325 | .192 |
| | F(3,45) = 0.864; *p* = .467; adjusted r² = -.009 | | | | |
| **Model 5** | | | | | |
| ODI | 5.935 | 42.718 | .023 | .139 | .890 |
| NDI | -59.598 | 44.016 | -.221 | -1.354 | .182 |
| | F(2,46) = 1.322; *p* = .277; adjusted r² = .013 | | | | |
| **Model 6** | | | | | |
| NDI | -62.537 | 38.197 | -.232 | -1.637 | .108 |
| | F(1,47) = 2.681; *p* = .108; adjusted r² = .034 | | | | |

*Note*: Abbreviations: HC, healthy controls, FA, fractional anisotropy, IVF, isotropic volume fraction, ODI, orientation dispersion index, NDI, neurite density index.

*S4. Detailed results of neuropsychological analyses*

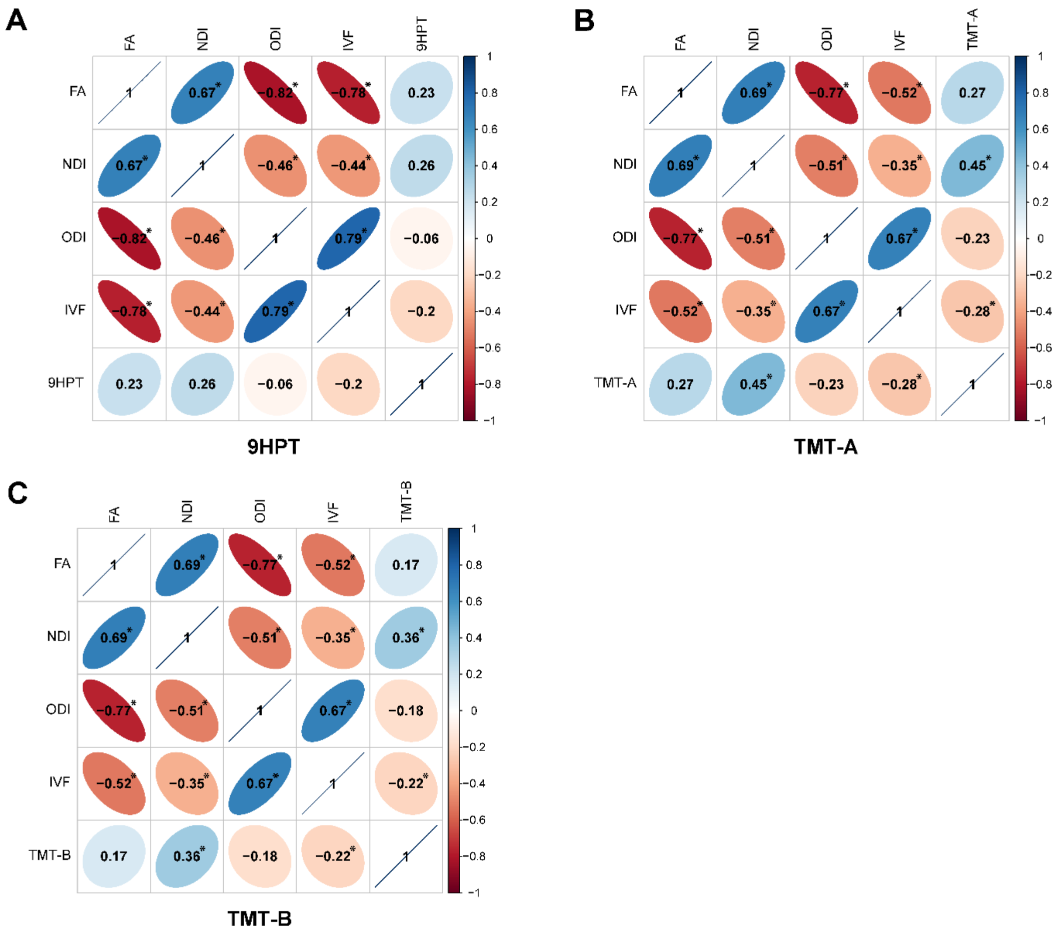

Figure S1. Correlation coefficients of the correlation between left M1 GM FA, NDI, ODI, IVF and (A) 9HPT,(B) TMT-A and (C) TMT-B in MS patients. * $p < .05$. Abbreviations: M1, primary motor cortex, GM, grey matter, FA, fractional anisotropy, NDI, neurite density index, ODI, orientation dispersion index, IVF, isotropic volume fraction, 9HPT, Nine Hole Peg Test, TMT-A, Trail Making Test part A, TMT-B, Trail Making Test part B, MS, multiple sclerosis.

Table S4. Results of hierarchical regression analysis of 9HPT

| | **Regression coefficient** | **Standard error** | **Beta coefficient** | **t statistic** | ***p* value** |
|---|---|---|---|---|---|
| **Model 1** | | | | | |
| Age | -.052 | .025 | -.424 | -2.074 | .049 |
| Gender | .198 | .643 | .069 | .307 | .761 |
| M1 lesion load | -.567 | .482 | -.265 | -1.178 | .251 |
| ODI | 24.673 | 17.248 | .595 | 1.430 | .166 |
| IVF | .880 | 5.522 | .062 | .159 | .875 |
| FA | 37.388 | 27.311 | .516 | 1.369 | .184 |
| NDI | 1.343 | 10.316 | .033 | .130 | .898 |

| | | | | | |
|---|---|---|---|---|---|
| | F(7,23) = 1.473; *p* = .226; adjusted r² = .099 | | | | |
| **Model 2** | | | | | |
| Age | -.051 | .024 | -.414 | -2.091 | .047 |
| M1 lesion load | -.559 | .472 | -.261 | -1.185 | .248 |
| ODI | 23.522 | 16.516 | .567 | 1.424 | .167 |
| IVF | .505 | 5.284 | .036 | .096 | .925 |
| FA | 35.866 | 26.347 | .495 | 1.361 | .186 |
| NDI | 2.297 | 9.650 | .056 | .238 | .814 |
| | F(6,24) = 1.769; *p* = .148; adjusted r² = .133 | | | | |
| **Model 3** | | | | | |
| Age | -.053 | .024 | -.436 | -2.196 | .038 |
| ODI | 17.785 | 15.918 | .429 | 1.117 | .274 |
| IVF | -.666 | 5.232 | -.047 | -.127 | .900 |
| FA | 20.717 | 23.223 | .286 | .892 | .381 |
| NDI | 7.536 | 8.648 | .184 | .871 | .392 |
| | F(5,25) = 1.813; *p* = .147; adjusted r² = .119 | | | | |
| **Model 4** | | | | | |
| Age | -.058 | .024 | -.476 | -2.426 | .022 |
| IVF | 3.217 | 3.930 | .227 | .818 | .421 |
| FA | 8.332 | 20.505 | .115 | .406 | .688 |
| NDI | 9.053 | 8.581 | .221 | 1.055 | .301 |
| | F(4,26) = 1.935; *p* = .134; adjusted r² = .111 | | | | |
| **Model 5** | | | | | |
| Age | -.050 | .022 | -.409 | -2.310 | .029 |
| FA | -2.550 | 15.514 | -.035 | -.164 | .871 |
| NDI | 9.124 | 8.528 | .223 | 1.070 | .294 |
| | F(3,27) = 2.386; *p* = .091; adjusted r² = .122 | | | | |
| **Model 6** | | | | | |
| Age | -.049 | .021 | -.401 | -2.385 | .024 |
| NDI | 8.324 | 6.879 | .204 | 1.210 | .236 |
| | F(2,28) = 3.694; *p* = .038; adjusted r² = .152 | | | | |
| **Model 7** | | | | | |
| Age | -.050 | .021 | -.409 | -2.415 | .022 |
| | F(1,29) = 5.831; *p* = .022; adjusted r² = .139 | | | | |

*Note*: Abbreviations: 9HPT, Nine Hole Peg Test, M1, primary motor cortex, ODI, orientation dispersion index, IVF, isotropic volume fraction, FA, fractional anisotropy, NDI, neurite density index.

Table S5. Results of hierarchical regression analysis of TMT-A

| | **Regression coefficient** | **Standard error** | **Beta coefficient** | **t statistic** | ***p* value** |
|---|---|---|---|---|---|
| **Model 1** | | | | | |
| Age | -.005 | .019 | -.046 | -.272 | .787 |
| Gender | .179 | .478 | .072 | .374 | .711 |
| M1 lesion load | -.469 | .369 | -.213 | -1.272 | .212 |
| ODI | 12.777 | 10.947 | .328 | 1.167 | .251 |
| FA | 1.106 | 16.458 | .018 | .067 | .947 |
| IVF | -3.705 | 3.227 | -.292 | -1.148 | .258 |
| NDI | 12.037 | 7.484 | .332 | 1.608 | .117 |
| | $F(7,34) = 1.923$; $p = .094$; adjusted $r^2 = .131$ | | | | |
| **Model 2** | | | | | |
| Gender | .150 | .460 | .060 | .325 | .747 |
| M1 lesion load | -.482 | .360 | -.219 | -1.338 | .189 |
| ODI | 13.119 | 10.738 | .337 | 1.222 | .230 |
| FA | 1.485 | 16.192 | .024 | .092 | .927 |
| IVF | -4.095 | 2.855 | -.323 | -1.434 | .160 |
| NDI | 11.727 | 7.304 | .324 | 1.606 | .117 |
| | $F(6,35) =2.289$; $p = .056$; adjusted $r^2 = .152$ | | | | |
| **Model 3** | | | | | |
| M1 lesion load | -.476 | .356 | -.216 | -1.337 | .189 |
| ODI | 12.656 | 10.517 | .325 | 1.203 | .236 |
| FA | .996 | 15.931 | .016 | .062 | .950 |
| IVF | -4.427 | 2.635 | -.349 | -1.680 | .101 |
| NDI | 12.373 | 6.945 | .342 | 1.782 | .083 |
| | $F(5,36) = 2.791$; $p = .030$; adjusted $r^2 = .172$ | | | | |
| **Model 4** | | | | | |
| ODI | 9.095 | 10.276 | .233 | .885 | .382 |
| FA | -5.886 | 15.229 | -.094 | -.387 | .701 |
| IVF | -4.883 | 2.639 | -.385 | -1.851 | .072 |

| | | | | | |
|---|---|---|---|---|---|
| NDI | 15.574 | 6.585 | .430 | 2.365 | .023 |
| | F(4,37) = 2.982; *p* = .031; adjusted r² = .156 | | | | |
| **Model 5** | | | | | |
| FA | -13.576 | 12.473 | -.217 | -1.088 | .283 |
| IVF | -3.464 | 2.090 | -.273 | -1.657 | .105 |
| NDI | 16.101 | 6.540 | .445 | 2.462 | .018 |
| | F(3,38) = 3.735; *p* = .019; adjusted r² = .160 | | | | |
| **Model 6** | | | | | |
| IVF | -2.491 | 1.893 | -.197 | -1.316 | .196 |
| NDI | 12.085 | 5.412 | .334 | 2.233 | .031 |
| | F(2,39) = 4.987; *p* = .012; adjusted r² = .156 | | | | |
| **Model 7** | | | | | |
| NDI | 14.565 | 5.117 | .402 | 2.846 | .007 |
| | F(1,40) = 8.102; *p* = .007; adjusted r² = .142 | | | | |

*Note*: Abbreviations: TMT-A, Trail Making Test part A, M1, primary motor cortex, ODI, orientation dispersion index, FA, fractional anisotropy, IVF, isotropic volume fraction, NDI, neurite density index.

Table S6. Results of hierarchical regression analysis of TMT-B

| | Regression coefficient | Standard error | Beta coefficient | t statistic | *p* value |
|---|---|---|---|---|---|
| **Model 1** | | | | | |
| Age | -.018 | .018 | -.173 | -.999 | .324 |
| Gender | .253 | .446 | .112 | .566 | .575 |
| M1 lesion load | -.325 | .344 | -.163 | -.944 | .351 |
| ODI | 4.940 | 10.224 | .140 | .483 | .632 |
| FA | -1.758 | 15.370 | -.031 | -.114 | .910 |
| IVF | -.388 | 3.014 | -.034 | -.129 | .898 |
| NDI | 11.692 | 6.990 | .357 | 1.673 | .103 |
| | F(7,36) = 1.485; *p* = .204; adjusted r² = .073 | | | | |
| **Model 2** | | | | | |
| Age | -.016 | .017 | -.151 | -.904 | .372 |
| M1 lesion load | -.320 | .341 | -.161 | -.939 | .354 |
| ODI | 4.345 | 10.075 | .123 | .431 | .669 |
| FA | -2.379 | 15.189 | -.042 | -.157 | .876 |

| | | | | | |
|---|---|---|---|---|---|
| IVF | -1.090 | 2.722 | -.095 | -.400 | .691 |
| NDI | 12.595 | 6.742 | .384 | 1.868 | .070 |
| | F(6,37) = 1.711; *p* = .146; adjusted r² = .090 | | | | |
| **Model 3** | | | | | |
| M1 lesion load | -.365 | .336 | -.183 | -1.085 | .285 |
| ODI | 5.639 | 9.949 | .160 | .567 | .574 |
| FA | -.960 | 15.072 | -.017 | -.064 | .950 |
| IVF | -2.065 | 2.492 | -.180 | -.829 | .412 |
| NDI | 11.289 | 6.570 | .345 | 1.718 | .094 |
| | F(5,38) = 1.899; *p* = .117; adjusted r² = .095 | | | | |
| **Model 4** | | | | | |
| ODI | 2.907 | 9.647 | .083 | .301 | .765 |
| FA | -6.241 | 14.296 | -.110 | -.437 | .665 |
| IVF | -2.415 | 2.477 | -.211 | -.975 | .335 |
| NDI | 13.745 | 6.182 | .420 | 2.223 | .032 |
| | F(4,39) = 2.070; *p* = .103; adjusted r² = .091 | | | | |
| **Model 5** | | | | | |
| FA | -8.699 | 11.607 | -.154 | -.749 | .458 |
| IVF | -1.962 | 1.945 | -.171 | -1.009 | .319 |
| NDI | 13.913 | 6.086 | .425 | 2.286 | .028 |
| | F(3,40) = 2.794; *p* = .053; adjusted r² = .111 | | | | |
| **Model 6** | | | | | |
| IVF | -1.339 | 1.749 | -.117 | -.766 | .448 |
| NDI | 11.340 | 4.998 | .346 | 2.269 | .029 |
| | F(2,41) = 3.952; *p* = .027; adjusted r² = .121 | | | | |
| **Model 7** | | | | | |
| NDI | 12.672 | 4.662 | .387 | 2.718 | .009 |
| | F(1,42) = 7.390; *p* = .009; adjusted r² = .129 | | | | |

*Note*: Abbreviations: TMT-B, Trail Making Test part B, M1, primary motor cortex, ODI, orientation dispersion index, FA, fractional anisotropy, IVF, isotropic volume fraction, NDI, neurite density index.

*S5. TBSS analysis applied to JHU atlas*

We computed the number and percentage of voxels showing group differences for each region of the JHU atlas for FA, NDI and ODI maps (Table S7). Fibers exhibiting group differences in a large part of the region in either FA, NDI or ODI or combinations of them included fornix, thalamic radiation, internal and external capsule, corpus callosum, corona radiata and sagittal stratum.

Table S7. Number and percentage of significant voxels in JHU regions for FA, NDI and ODI. The last column contains the total number of voxels of the mean skeleton in each region. Contrast HC > MS for FA and NDI, and MS > HC for ODI, sorted decreasingly by the percentage of voxels showing significant group differences of each region, summed over FA, NDI and ODI

| JHU label | FA (%) | NDI (%) | ODI (%) | Total |
|---|---|---|---|---|
| Left fornix (cres) / stria terminalis | 323 (28.7) | 268 (23.8) | 297 (26.4) | 1125 |
| Right fornix (cres) / stria terminalis | 296 (26.3) | 187 (16.6) | 294 (26.4) | 1124 |
| Left posterior thalamic radiation | 1128 (28.4) | 1144 (28.8) | 401 (10.1) | 3978 |
| Left retrolenticular part of internal capsule | 614 (24.9) | 668 (27.1) | 224 (9.1) | 2469 |
| Right posterior thalamic radiation | 1136 (28.6) | 1168 (29.4) | | 3972 |
| Body of corpus callosum | 3076 (22.3) | 3003 (21.9) | 1666 (12.2) | 13711 |
| Right sagittal stratum | 530 (23.8) | 586 (26.3) | | 2228 |
| Genu of corpus callosum | 1734 (19.6) | 1733 (19.6) | 860 (9.7) | 8851 |
| Left sagittal stratum | 450 (10.2) | 476 (21.3) | 89 (4) | 2231 |
| Left external capsule | 879 (15.7) | 805 (14.4) | 775 (13.9) | 5587 |
| Left anterior corona radiata | 1145 (16.7) | 1505 (22) | 138 (2) | 6852 |
| Right retrolenticular part of internal capsule | 445 (17.7) | 577 (22.9) | 1 (0.04) | 2515 |
| Right posterior corona radiata | 690 (18.5) | 759 (20.4) | 20 (0.5) | 3728 |
| Splenium of corpus callosum | 2295 (18) | 2359 (18.5) | 120 (0.9) | 12729 |
| Left superior longitudinal fasciculus | 968 (14.7) | 1402 (21.2) | 3 (0.04) | 6605 |
| Fornix (column and body) | 80 (12.1) | 70 (10.6) | 86 (13.1) | 659 |
| Right anterior corona radiata | 714 (10.4) | 1449 (21.2) | 251 (3.7) | 6849 |
| Right superior longitudinal fasciculus | 874 (13.2) | 1411 (21.4) | | 6607 |
| Left posterior corona radiata | 573 (15.4) | 684 (18.4) | 21 (0.6) | 3714 |
| Left superior corona radiata | 901 (12) | 1226 (16.3) | 202 (2.7) | 7508 |
| Right superior corona radiata | 775 (10.3) | 1345 (17.9) | 198 (2.6) | 7500 |
| Right cerebral peduncle | 359 (15.8) | | 282 (12.4) | 2278 |
| Left superior fronto-occipital fasciculus | 3 (0.6) | 106 (20.9) | 29 (5.7) | 507 |

| | | | | |
|---|---|---|---|---|
| Right anterior limb of internal capsule | 216 (6.9) | 88 (2.8) | 532 (17) | 3138 |
| Right superior fronto-occipital fasciculus | | 73 (14.4) | 45 (8.9) | 507 |
| Right cingulum (hippocampus) | 103 (8) | 175 (14.2) | | 1236 |
| Left posterior limb of internal capsule | 12 (0.3) | 407 (10.8) | 373 (9.9) | 3752 |
| Left uncinate fasciculus | 33 (8.8) | 22 (2.9) | 23 (6.1) | 376 |
| Right cingulum (cingulate gyrus) | 120 (5.1) | 323 (13.8) | | 2342 |
| Left cerebral peduncle | | 5 (0.2) | 421 (18.5) | 2278 |
| Left anterior limb of internal capsule | 2 (0.1) | 95 (3.1) | 438 (14.5) | 3018 |
| Left cingulum (hippocampus) | | 200 (17.3) | | 1155 |
| Right external capsule | 378 (6.7) | 558 (9.9) | | 5611 |
| Left cingulum (cingulate gyrus) | 42 (1.5) | 390 (14.2) | | 2751 |
| Right posterior limb of internal capsule | 307 (8.2) | 89 (2.4) | 160 (4.3) | 3754 |
| Right tapetum | 36 (6) | 36 (6) | | 596 |
| Right corticospinal tract | 29 (2.1) | | 133 (9.8) | 1362 |
| Right superior cerebellar peduncle | | | 81 (8.2) | 992 |
| Right inferior cerebellar peduncle | | | 79 (8.2) | 968 |
| Right medial lemniscus | | | 28 (4.1) | 690 |
| Middle cerebellar peduncle | | | 595 (3.8) | 15644 |
| Right uncinate fasciculus | 2 (0.5) | 8 (2.1) | | 380 |
| Left tapetum | 4 (0.7) | 4 (0.7) | | 600 |
| Left corticospinal tract | | | 9 (0.7) | 1370 |
| Pontine crossing tract | | | 7 (0.5) | 1500 |
| **TOTAL** | **21272 (12.7)** | **25404 (15.2)** | **8881 (5.3)** | **167347** |

*Note:* Abbreviations: FA, fractional anisotropy, NDI, neurite density index, ODI, orientation dispersion index, HC, healthy control, MS, multiple sclerosis.

Again, we disentangled if one of the parameters alone or if combinations of FA, NDI and ODI contributed to observed group differences. The percentage of all single parameters and their combinations are depicted for each JHU region is presented in Figure S4A, and averages over all regions is depicted in Figure S4B. Neurite density exclusively (49%) and in combination with FA (29%) accounted for the largest amount of differences, followed by a combination of NDI, ODI and FA (9%) and ODI alone (6%).

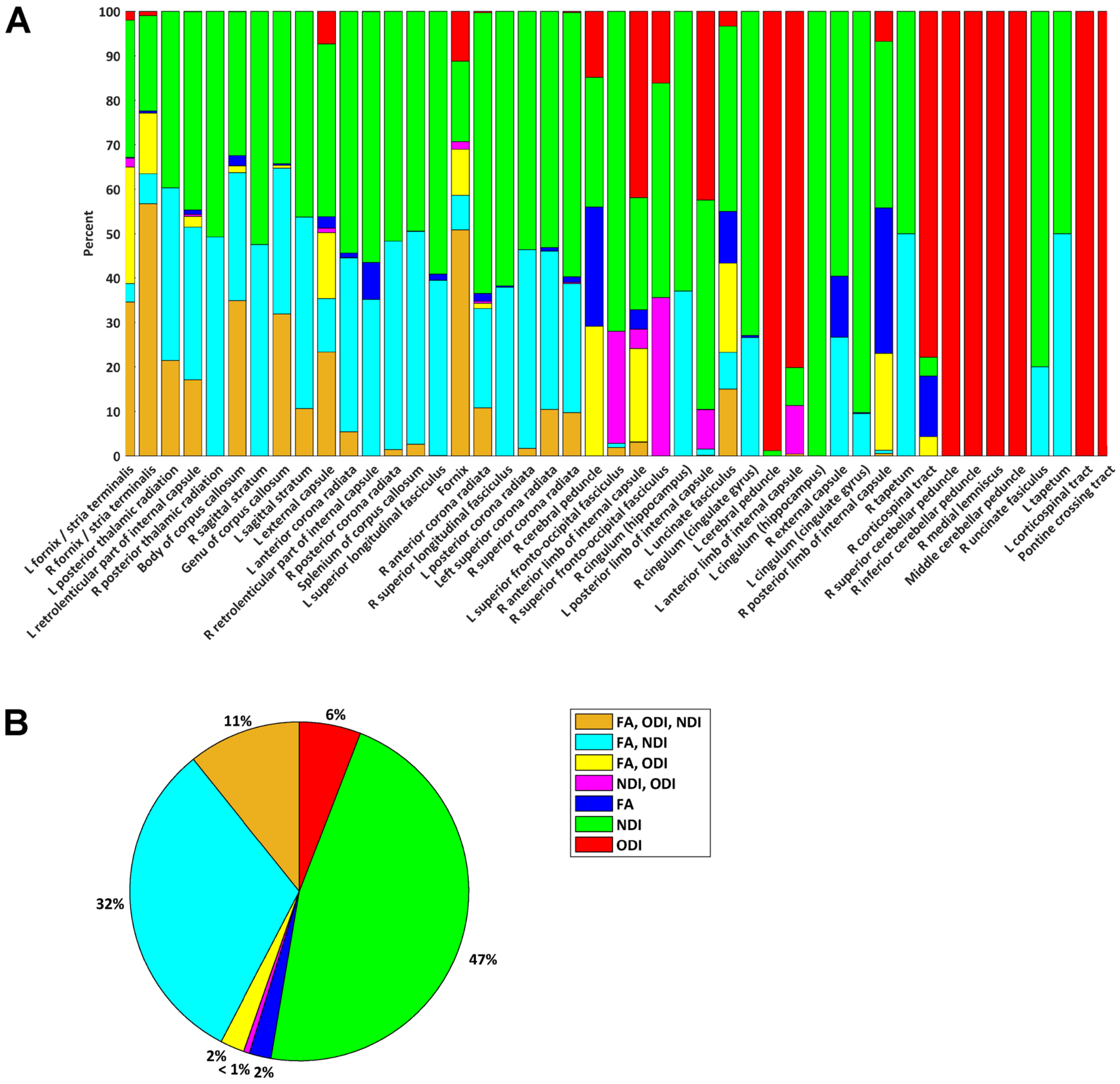

Figure S2. Percentage of FA, NDI and ODI separately or combined showing group differences in the tract skeleton in intersection with JHU regions (A). Percentage averaged over regions with any significant group difference (B). Abbreviations: L, left, R, right, FA, fractional anisotropy, NDI, neurite density index, ODI, orientation dispersion index
